# To RAG, or Not to RAG? A Comparative Evaluation of Retrieval-Augmented Generation for ICD Coding of German Tumor Diagnoses

**DOI:** 10.64898/2026.05.27.26353695

**Authors:** Fatma Alickovic, Stefan Lenz, Arsenij Ustjanzew, Lakisha Ortiz Rosario, Georg Vollmar, Thomas Kindler, Torsten Panholzer

## Abstract

**Introduction:** Coding tumor diagnoses from free-text clinical documentation currently requires substantial manual effort. Promising approaches for automating this process include large language models (LLMs), embedding models, and retrieval-augmented generation (RAG). While previous studies often focus on a single method, we directly compare these approaches on a real-world dataset of tumor diagnosis descriptions to assess their strengths and limitations.

**Methods:** We evaluated nine different embedding models using similarity search and embedding-based classification, as well as LLM-based coding, with and without RAG, on a real-world dataset of 2,024 unique German tumor diagnosis descriptions labeled with ICD-10 and ICD-O topography codes. The retrieval knowledge base was constructed exclusively from standardized Alpha-ID, ICD-10-GM, and ICD-O-3 classifications. Performance was assessed for exact (full-code) and partial (three-character) code prediction. For RAG, we evaluated base and fine-tuned versions of Llama 3.1 8B and Llama 3.3 70B.

**Results:** Qwen3-Embedding-8B, the largest embedding model, yielded the best results. It achieved 47.8% exact-match and 72.1% partial-match accuracy for ICD-10 coding with classification, and 42.7% exact-match and 73.5% partial-match accuracy for ICD-O coding with similarity search. The other embedding models, including medically specialized ones, showed varied but lower performance. RAG improved base LLM performance and outperformed embedding-based approaches on partial-match accuracy (80.6% partial-match accuracy for ICD-10 and 75.0% for ICD-O with Llama 3.3 70B), but not on exact-match accuracy.

**Conclusion:** A direct comparison with embedding-based approaches is essential to determine whether the additional effort of RAG is justified. The strong variation in performance also highlights the importance of model selection. Further advances in embedding-based methods, potentially supported by larger and more diverse training data, may offer a promising direction for future work.

## Introduction

The clinical coding of tumor diagnoses is foundational for tumor documentation and cancer registry reporting ^1^. In Germany, this process commonly involves two complementary classification systems: the German Modification of the International Classification of Diseases, 10th Revision (ICD-10-GM) for diagnosis coding ^2^, and the International Classification of Diseases for Oncology, Third Edition (ICD-O-3), which captures oncology-specific information through a topography code indicating tumor location by anatomical site and a morphology code providing histological detail of the tumor ^3^.

In routine clinical documentation intended for communication between physicians, standardized diagnostic codes are often omitted, and diagnoses are instead recorded as free-text descriptions. For secondary use of clinical data, particularly statistical reporting and cancer research, these descriptions must be translated into standardized classification systems. This translation is typically performed by tumor registrars, who review clinical reports, extract the relevant diagnostic information, and assign the corresponding codes in dedicated tumor documentation databases. Automating this process is challenging because diagnosis descriptions exhibit substantial lexical and stylistic variation, which limits rule-based or dictionary-based matching approaches. Heterogeneous clinical jargon, variable levels of granularity, frequent abbreviations, and typographical errors further exacerbate the problem ^4^. As a result, fully automated tumor coding has remained difficult to achieve in practice.

Large Language Models (LLMs) offer a promising solution, as they can robustly interpret heterogeneous free-text inputs and have been pretrained on large corpora that support generalization across linguistic variation. Several approaches have been explored for mapping diagnosis descriptions to classification codes with LLMs ^4–6^. One approach uses the models to create high-dimensional vector representations, so-called embedding vectors, that capture the meaning of text in a multidimensional vector space. Based on these embedding vectors, coding can either be performed by training a classifier for multi-label classification, or by similarity search in a vector space of embeddings. Based on the distances of the embedding vectors, texts with similar meaning can be identified. Many embedding models are based on the BERT architecture ^7,8^. Newer generative LLMs, which can respond with textual answers, allow simple, text-based prompting with questions as another way to map texts to codes ^9^. An approach that combines both the use of embedding vectors and generative LLMs is retrieval-augmented generation (RAG), which has also been explored for ICD-10 coding ^6^. RAG combines a database of embeddings as a knowledge base for retrieving relevant information with the ability of LLMs to integrate information and respond to text-based queries.

The objective of the present study is to provide a direct, side-by-side comparison of the components of the RAG method to systematically assess the performance of ICD coding of German tumor diagnoses using embedding models and generative AI on a single, real-world test dataset. For building the knowledge base of the models, we use only standardized classifications to avoid site-specific bias in the results. Particularly advantageous here is the German version of the ICD-10, which comes with an alphabetical index that captures some lexical variation as well as different granularity of the diagnosis descriptions ^2^. Based on this data, we evaluate embedding-based similarity search and embedding-based classification across multiple embedding models of varying sizes. For the generative LLMs, we use base and fine-tuned versions from the Llama 3 model family ^10^ at multiple scales. Together, these experiments provide an empirical basis for selecting LLM-based coding strategies for German tumor documentation.

## Methods

This study was reported in accordance with the TRIPOD-LLM statement ^11^, a reporting guideline for research involving large language models.

### Construction of the knowledge base

To construct a standardized and comprehensive knowledge base for RAG prompting, publicly accessible German medical classifications were utilized. These resources offer consistent terminology and broad coverage of the diagnostic codes routinely used in tumor documentation. The following ICD classifications were utilized to create the knowledge base:

- Alpha-ID: Alphabetical index of the German version of the International Classification of Diseases (ICD-10-GM), 2024 edition, containing a diverse set of diagnostic descriptions to ICD-10 codes ^12^.
- ICD-10-GM: German version of the International Classification of Diseases, 2024 edition, providing standardized diagnostic code definitions ^13^.
- ICD-O-3: International Classification of Diseases for Oncology, German version, second revision, 2019, including topography and morphology specifications for tumor diagnoses ^3^.

The study focused exclusively on oncological codes from Chapter II (Neoplasms; C00–D48) of the Alpha-ID and ICD-10-GM catalogues, consistent with its emphasis on tumor diagnosis coding. Codes C77–C79, which denote secondary malignant neoplasms (metastases), were excluded, as they are not used for tumor documentation, where only the primary tumor diagnoses are coded ^9^.

To increase lexical coverage for ICD-O coding, a mapping table between the Alpha-ID dataset and ICD-O-3 was utilized, which has been developed by an expert committee for cancer registration in Germany ^14^. This mapping table corresponds to ICD-O-3.2 (2019 revision). For all one-to-one mappings between ICD-10 codes and ICD-O-3, the diagnosis descriptions in the Alpha-ID could be mapped to ICD-O-3 codes via the mapping table.

Ambiguous mappings were resolved by combining the Alpha-ID diagnosis text with the corresponding ICD-O topography description. When localization information from ICD-O-3 was incorporated into the final diagnosis text, non-specific expressions in the original ICD-10 description (e.g., *“other site”*; German: *„Sonstige Lokalisation”*) were removed if the anatomical site was explicitly defined through the ICD-O-3 mapping. For example, ICD-10 code D18.08 (“Hämangiom: Sonstige Lokalisation” [hemangioma: other site]) was mapped to the ICD-O-3 topography code C76 (“Kopf, Gesicht oder Hals o.n.A.” [head, face or neck, NOS]). Because the ICD-O-3 description provides a specific anatomical region, the non-specific designation “Sonstige Lokalisation” was omitted, resulting in the consolidated diagnosis text: “Hämangiom (Kopf, Gesicht oder Hals ohne nähere Angabe)” [hemangioma (head, face or neck, not otherwise specified)]. Additionally, abbreviations were expanded to ensure consistent and unambiguous anatomical terminology. In the following, this mapped dataset is referred to as the ICD-O-3 Lexically Enriched Dataset (ICD-O-3-LE), an overview of its entries is provided in Table 1. As can be seen there, two separate knowledge bases were used for ICD-10 and ICD-O coding. All diagnosis descriptions with their corresponding codes were used as distinct items in the knowledge base.

**Table 1.** Overview of the datasets used in this study, including metadata and intended purpose, with key characteristics such as dataset version, size, and unique code count.

| Dataset | Year/<br>Version | No. of<br>entries | No. of unique<br>codes | Purpose |
| --- | --- | --- | --- | --- |
| Alpha-ID | 2024 | 12.021 | 776 | Knowledge base for ICD-10 coding |
| ICD-10-GM | 2024 | 859 | 859 | Standardized diagnosis descriptions for RAG prompts |
| ICD-O-3-LE | 2024/2019 | 8981 | 615 (ICD-10)<br>326 (ICD-O) | Knowledge base for ICD-O coding |
| Test dataset | 2023–2024 | 2.024 | 238 (ICD-10)<br>183 (ICD-O) | Evaluation of model performance |

### Test dataset

For model evaluation, a real-world dataset extracted from the local tumor documentation system on 27 June 2024 was employed. To ensure compatibility with the current ICD-10-GM version and the Alpha-ID classification, only cases from 2023–2024 were included, and duplicate entries were removed. The resulting test set comprised 2,024 unique tumor diagnosis descriptions in free text, each of them annotated with an ICD-10 code and an ICD-O topography code. These data were previously used as a test dataset in the study evaluating instruction-based fine-tuning of LLMs for ICD-10 and ICD-O-3 topography coding ^5^.

To improve retrieval performance, acronyms in the diagnosis descriptions were expanded. This was done via a rule-based and embedding-supported similarity approach: A list of medical acronyms and their corresponding long forms ^15^ was compiled and supplemented with additional, manually defined German clinical abbreviations. Acronyms with only one clear meaning have been replaced directly with the corresponding long form. If multiple candidate expansions were available, the correct expansion was selected using a sentence-embedding similarity approach and cosine similarity. Specifically, the diagnosis text and candidate long forms were encoded using the jina-embeddings-v2-base-de model ^16^, implemented via the Sentence Transformers Python package (version 4.48.0) ^17^.-

The selected expansion was appended to the original diagnosis text in brackets to preserve the original terminology while enriching semantic information. For example, the acronym “CUP” was extended to “CUP (cancer of unknown primary site)”.

In the resulting test dataset, dermatological diagnoses were particularly prevalent. Specifically, ICD-10 codes C44.3 and C43.5 were the most frequent, comprising 8.25% and 4.3% of all cases, respectively, as shown in Figure 1.

**Figure 1.**
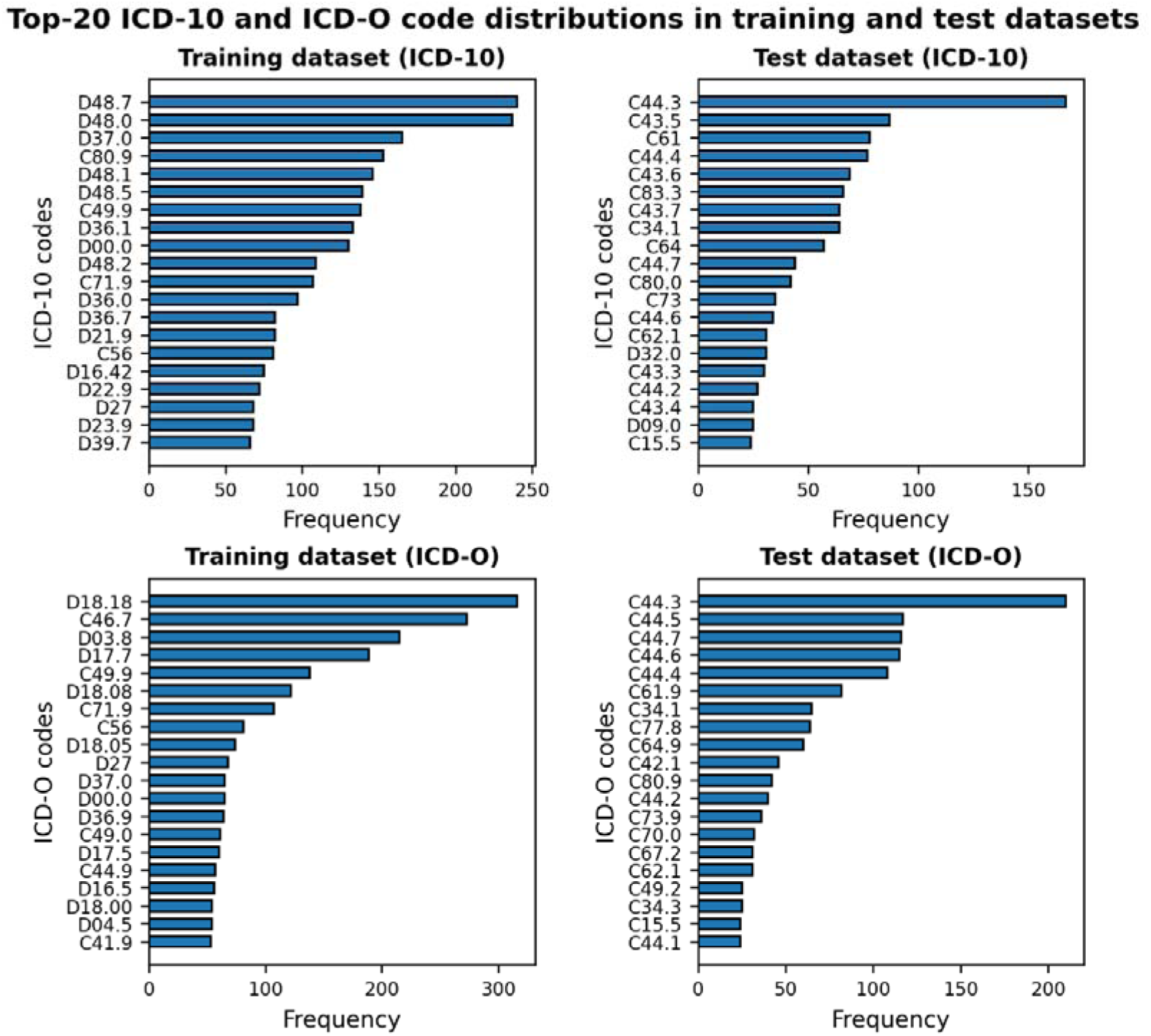
Class distribution of the training datasets (Alpha-ID and ICD-O-3-LE classifications) compared with the test dataset

The free-text tumor descriptions in the test dataset originated from more comprehensive routine clinical documentation, and therefore did not always provide sufficient information for unambiguous coding on their own. A data quality analysis in our previous work ^5^ identified unspecified tumor behavior and missing anatomical site information as limiting factors for reliable ICD-10 and ICD-O assignment. Consequently, the maximum achievable accuracy was limited by data quality, necessitating consideration of performance upper bounds for ICD-10 and ICD-O-3 coding. For exact code derivation (full match), the upper bound was estimated at 70% (95% CI: 60–79%) for ICD-10 and 65% (95% CI: 55–75%) for ICD-O-3. For partial matches (three-character codes only), the estimated ceilings were 89% (95% CI: 81–94%) for ICD-10 and 91% (95% CI: 84–96%) for ICD-O-3. These values served as reference limits in the subsequent evaluation, indicating the maximum attainable performance under the given data conditions.

### Evaluation metrics

To enable a detailed assessment of LLM performance, accuracy, weighted F1 score, and macro F1 score were computed for the predicted code per diagnosis description, using exact match and partial match criteria. Under exact match, a prediction was considered correct only if the predicted ICD code string was identical to the reference code. Partial match credited agreement at the three-character category level, allowing deviations at more specific subcategory levels.

In addition to accuracy, the weighted F1 score was reported to facilitate comparison with prior automated clinical ICD coding studies ^4^. The weighted F1 score combines precision and recall into a single measure and averages per-class F1 values, weighted by class support (the number of actual instances) in the test set, thereby assigning greater influence to more frequent codes. Moreover, we report the macro F1 score as the arithmetic mean of per-class F1 scores, treating all codes equally regardless of frequency. As a result, macro F1 is more sensitive to performance on rare diagnoses and provides a complementary perspective on model robustness across the full code spectrum, compared to weighted F1 score and accuracy ^18^.

### Overview of RAG pipeline

Our RAG pipeline integrated embedding-based retrieval with LLMs. Retrieved ICD codes were incorporated into a structured RAG prompt, which was subsequently processed by the LLM to generate the final prediction (see Figure 2).

**Figure 2.**
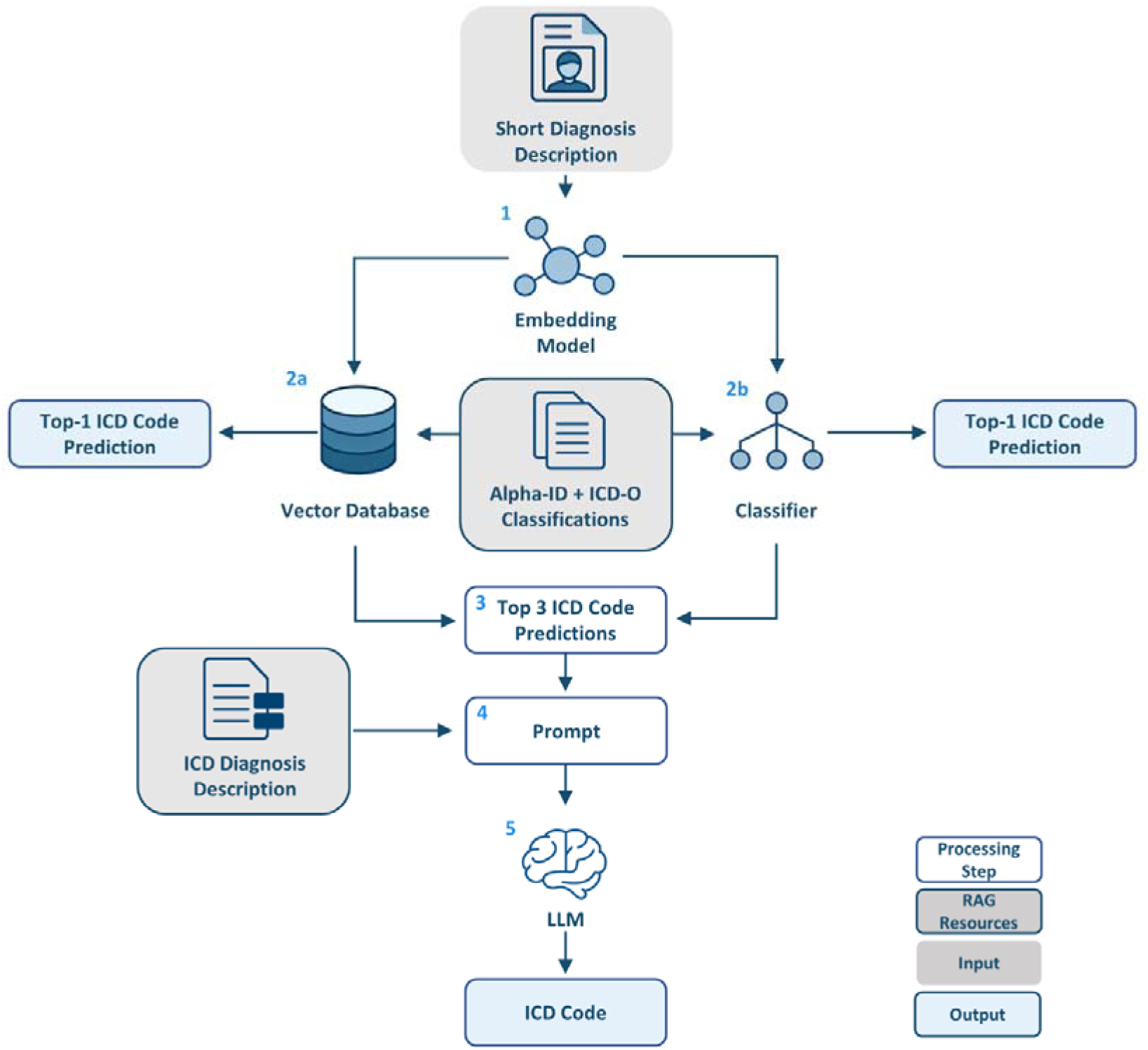
Overview of the used RAG pipeline for ICD code prediction. The RAG examples are identified using either semantic retrieval from a vector database or supervised classification via a neural network trained on the embeddings and the corresponding code classes. The two alternative approaches are evaluated on their own using top-1 ICD code predictions, while the top-3 candidates are incorporated into a structured prompt to support LLM-based inference and generate the final ICD code prediction via RAG.

First, short, free-text diagnosis descriptions were encoded into dense vector representations using pre-trained embedding models (Figure 2, step 1). These embeddings formed the basis for two alternative prediction strategies. In the first approach, semantic similarity search was conducted against vectorized entries from the Alpha-ID and ICD-O classification systems stored in a vector database (step 2a). The most similar codes were retrieved using cosine similarity, yielding ranked top-1 candidate predictions. The embedding models included larger pre-trained language models, as well as Word2Vec, a simpler neural embedding method. The latter was used as a baseline due to its previously demonstrated strong performance in ICD-10-related semantic retrieval for German clinical text ^19^.

In the second approach, embeddings were used as inputs for a neural network, which we refer to as the classification head here, to directly predict ICD codes, yielding a top-1 prediction (step 2b). The top-1 predictions obtained from the classifier and the semantic similarity search were compared to determine the better-performing method for improving prompt quality.

Within the RAG framework, up to three retrieved ICD codes and their corresponding diagnosis descriptions from the ICD-10-GM or ICD-O-3 datasets were retrieved (step 3) and integrated into a structured prompt together with the original medical query (step 4). Here, the standardized descriptions for the codes from the ICD-10 and ICD-O classification were used instead of the Alpha-ID and ICD-O-3-LE texts. This choice was made to avoid overly specialized descriptions. If duplicated codes were predicted, only one instance was kept. In each retrieved example, the prompt format consisted of an instruction followed by the specific question, reflecting the question format used during LLM training. An example is shown in the Supplemental Material. The resulting prompt supplied the LLM with contextualized candidate information, enabling it to generate the final ICD code prediction (step 5).

### Implementation of similarity search and selection of embedding models

To implement embedding-based similarity search, the ChromaDB ^20^ vector database was employed as the retrieval backend. The ICD-10 and ICD-O-3 classifications served as the primary knowledge sources and were ingested into the database. Each Alpha-ID entry was encoded using the selected embedding models and stored in dedicated ChromaDB collections to enable efficient nearest-neighbor retrieval.

The Word2Vec model was trained using the *bmschmidt/wordVectors* implementation in R ^21^, with Skip-Gram objective and a context window size of 10 tokens, 30 training iterations, and 10 negative samples per word. Nearest-neighbor retrieval was performed using cosine similarity, with Jaro–Winkler similarity additionally evaluated for lexical matching ^22^.

For each query, up to three Alpha-ID entries with the highest cosine similarity scores were retrieved and used as candidate labels for subsequent prompt construction within the RAG framework.

The embedding models underlying this retrieval step were selected based on four criteria: language coverage, MTEB performance ^23^, domain specialization, and embedding dimensionality.

All selected models support the German language, which is essential for accurately capturing domain-specific semantic and syntactic patterns in German clinical texts.

In order to establish a robust baseline, a selection of embedding models was made based on their performance on the MTEB, a comprehensive evaluation framework designed to assess the quality of text embedding models across a variety of downstream tasks, including semantic textual similarity (STS), retrieval, classification, clustering, and reranking ^23^. The STS task evaluates an embedding model’s ability to capture fine-grained semantic similarity between text pairs. Qwen3-Embedding-8B, me5, and jina-embeddings-v3 were therefore selected, achieving Spearman’s rank correlation coefficients of 81.08, 76.81, and 77.31, respectively (benchmark scores retrieved in November 2025).

Domain specialization was further considered by including medically pretrained models, such as MedBERT.de and SapBERT, which serve as reference points for assessing the impact of clinical domain knowledge relative to general-purpose embedding models.

The selected models span embedding dimensionalities from 768 to 4096 and parameter sizes ranging from approximately 100 million to 8 billion. This variation enables analysis of the relationship between representational capacity, computational complexity, and ICD code prediction performance. An overview of the models and their characteristics can be found in Table 2.

**Table 2.** Overview of the embedding models, including model size (number of parameters), number of embedding dimensions, medical domain specialization, and release year. The models are ordered by the number of model parameters.

| Embedding model | Number of parameters | Embedding dimension | Medical domain | Release year | Model Card |
| --- | --- | --- | --- | --- | --- |
| Qwen3-Embedding-8B | 8B | 4096 | No | 2025 | <sup>24</sup> |
| jina-embeddings-v3 | 572M | 1024 | No | 2024 | <sup>25</sup> |
| mE5 | 560M | 1024 | No | 2024 | <sup>26</sup> |
| deepset-mxbai-embed | 487M | 1024 | No | 2024 | <sup>27</sup> |
| Nomic-Embed-v2 | 475M | 768 | No | 2025 | <sup>28</sup> |
| SapBERT | 278M | 1024 | Yes | 2024 | <sup>29</sup> |
| jina-embeddings-v2 | 161M | 768 | No | 2024 | <sup>16</sup> |
| ModernBERT | 149M | 768 | No | 2025 | <sup>30</sup> |
| medBERT.de | 109M | 768 | Yes | 2023 | <sup>31</sup> |

### Classification head architecture

As an alternative to the vector database-based similarity search, supervised classification was implemented, in which the selected pretrained sentence embedding models (Table 2) were coupled with a classification head. The classifier was trained on embeddings of textual diagnosis descriptions from the Alpha-ID dataset as input features and the corresponding codes as labels.

The classifier architecture comprised a fully connected hidden layer with 256 units, applied to the sentence embeddings. Batch normalization, ReLU activation, and dropout (rate = 0.2) were applied to this layer to stabilize training and mitigate overfitting. A final linear layer projected the hidden representations onto the ICD code label space. During training, weighted cross-entropy loss was used to compare the resulting class scores with the true labels.

To compensate for the imbalance among ICD classes in the training dataset, we employed a class-weighted cross-entropy loss. Class weights were computed inversely proportional to the frequency of each code and normalized relative to the most frequent class, thereby assigning higher penalties to misclassifications of rare diagnoses ^32,33^. Model optimization was performed using the AdamW optimizer ^34^, with identical hyperparameter settings applied across all embedding models to ensure fair and reproducible comparisons. Training parameters, including the learning rate, batch size, and weight decay, were optimized using the Optuna hyperparameter optimization framework (19). Training was performed with a batch size of 16, a learning rate of 2.17 × 10⁻⁴, and a weight decay of 3.44 × 10⁻³ for ICD-10 coding. The training parameters for ICD-O coding are presented in Table S2 of the Supplemental Material.

The final models were trained on a single NVIDIA A40 GPU with 48 GB of VRAM, using CUDA 12.2. Due to differences in encoder architecture size, total training time varied substantially across models, ranging from approximately 7.7 minutes for medBERT.de (109M parameters) to 5.2 hours for Qwen3-Embedding-8B (8B parameters) in ICD-10 coding.

### Large Language Models

Base and fine-tuned variants of Llama 3.1 8B ^35,36^ and Llama 3.3 70B ^37,38^ were evaluated. The fine-tuned models were derived from prior work, in which eight LLMs were instruction-tuned on publicly available clinical datasets for ICD coding, including the Alpha-ID dataset and ICD-O-3 ^5^. Llama 3.3 70B was selected as the best-performing model based on accuracy in the prior study, while Llama 3.1 8B was included to examine the impact of model size within the same model family. Fine-tuned versions of the models from the previous study were also evaluated. Parameter-Efficient Fine-Tuning (PEFT) ^39^ had been performed using Low-Rank Adaptation (LoRA) ^40^.

To investigate the LLM performance in different scenarios, three prompting setups were evaluated: 1) zero-shot prompting, reflecting the baseline LLM performance; 2) few-shot prompting with randomly selected examples from the ICD-10-GM and ICD-O-3-LE datasets; and 3) RAG prompting, where the prompt is augmented with the top candidates produced by the retrieval methods. Comparing few-shot prompting with random examples against RAG with retrieved examples allowed to distinguish gains from the prompting format itself from gains due to the retrieved information.

During inference, the temperature was set to 0, and all other generation parameters were left at their default values to ensure deterministic outputs. In addition, the maximum number of generated tokens was set to 20. Inference was conducted on three NVIDIA A40 GPUs, with 48 GB VRAM each, using CUDA 12.2. Due to hardware memory constraints, the 70B model was loaded using 4-bit quantization during inference, whereas the 8B model was executed without quantization.

## Results

### Performance comparison of semantic search and its classifier-enhanced variant

Figure 3 shows a side-by-side comparison of the performance of the different embedding models with respect to top-1 predictions of ICD-10 codes on the test data set. The trained classifier head yielded higher accuracy and weighted F1 scores for nearly all embedding models, compared to the semantic similarity search. The differences were especially notable regarding the partial accuracy and less pronounced regarding the exact accuracy. The only exception to this pattern was SapBERT, which was the only model that performed considerably worse with the classification layer. This finding demonstrates that supervised decision layers could leverage embedding features beyond nearest-neighbor retrieval, particularly in cases where exact matches are being compared.

**Figure 3.**
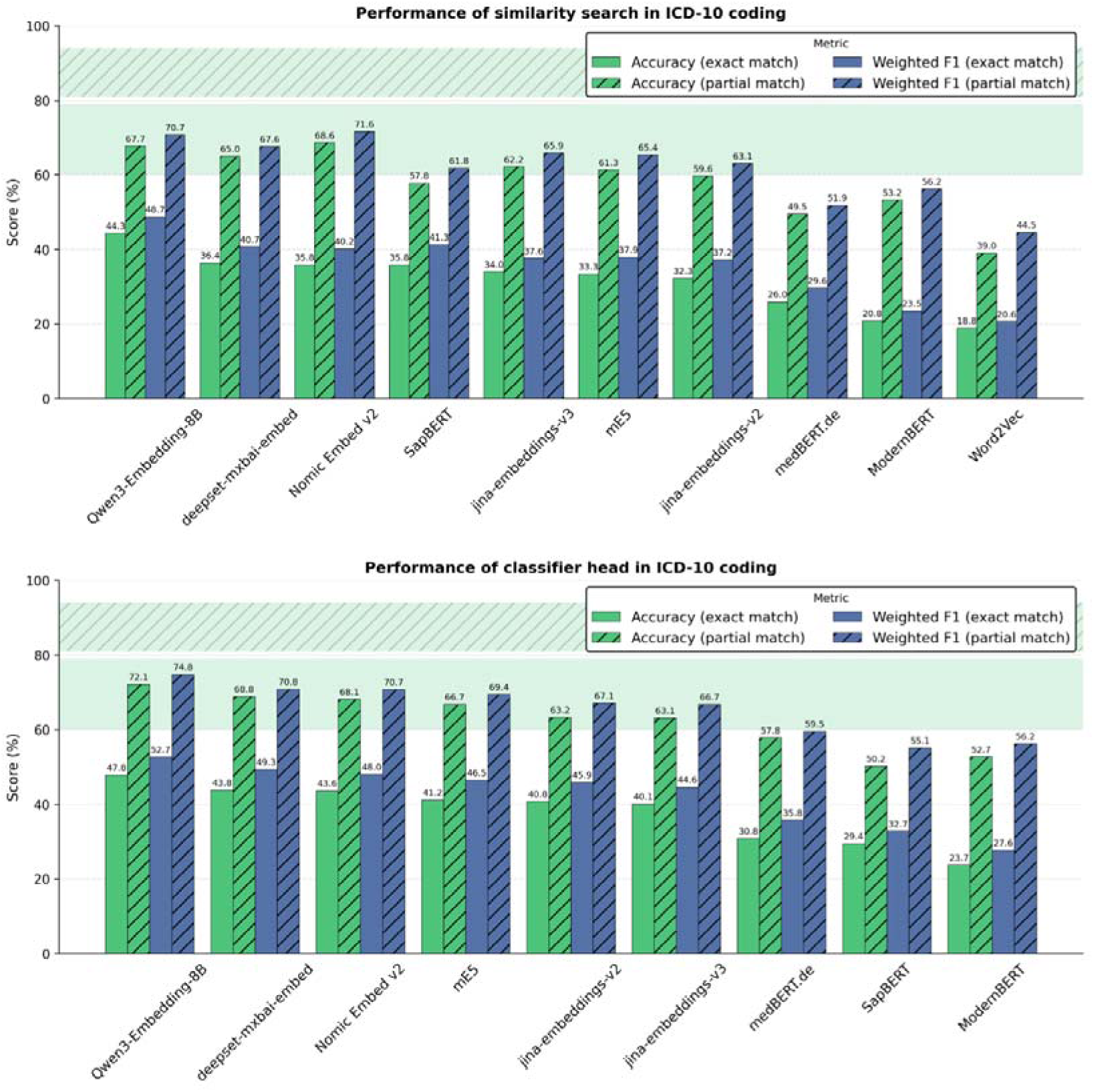
Comparison of similarity search (top) and classification (bottom) using embedding models, ICD-10 coding, with respect to accuracy and weighted F1 under exact- and partial-match evaluation on the test dataset. Embedding models are ordered by decreasing exact-match accuracy. The green bands show the 95% confidence interval for the maximum attainable accuracy according to the data quality analysis. The upper dashed band indicates the partial match accuracy limit and the lower band the exact match accuracy limit.

In both approaches, the largest model, Qwen3-Embedding-8B, achieved the strongest overall performance. Its values for the exact accuracy were 44.3% and 47.8% for similarity search and classification, and for the partial accuracy 67.7% and 72.1%, respectively. The smallest model, Word2Vec, markedly underperformed newer and larger transformer-based embedding models across all evaluation metrics, which is in line with the general observation that models with more parameters tended to perform better. Notably, domain-specific medical embeddings did not uniformly translate into superior performance, as the two models trained for the medical domain, SapBERT and medBERT did not outperform the other models comparable with their size.

Across all embedding models, partial-match metrics consistently exceeded exact-match metrics by a large margin, indicating that most errors originate from fine-grained code disambiguation rather than complete semantic mismatch.

While the ICD-10 coding system demonstrated benefits from the incorporation of a classifier head, particularly in the context of exact matches, the ICD-O coding system demonstrated the opposite trend, with retrieval-only similarity search outperforming the classifier-augmented variant (see Figure 4). Across all embedding models, the classifier head was associated with lower accuracy and weighted F1 scores, with the most pronounced difference observed for partial-match evaluation. Although Qwen3-Embedding-8B demonstrated the strongest overall performance in both setups, the values for the exact accuracy were lower, from 42.7% to 33.8%, and its partial accuracy decreased from 73.5% to 60.9% in the classifier-head configuration.

**Figure 4.**
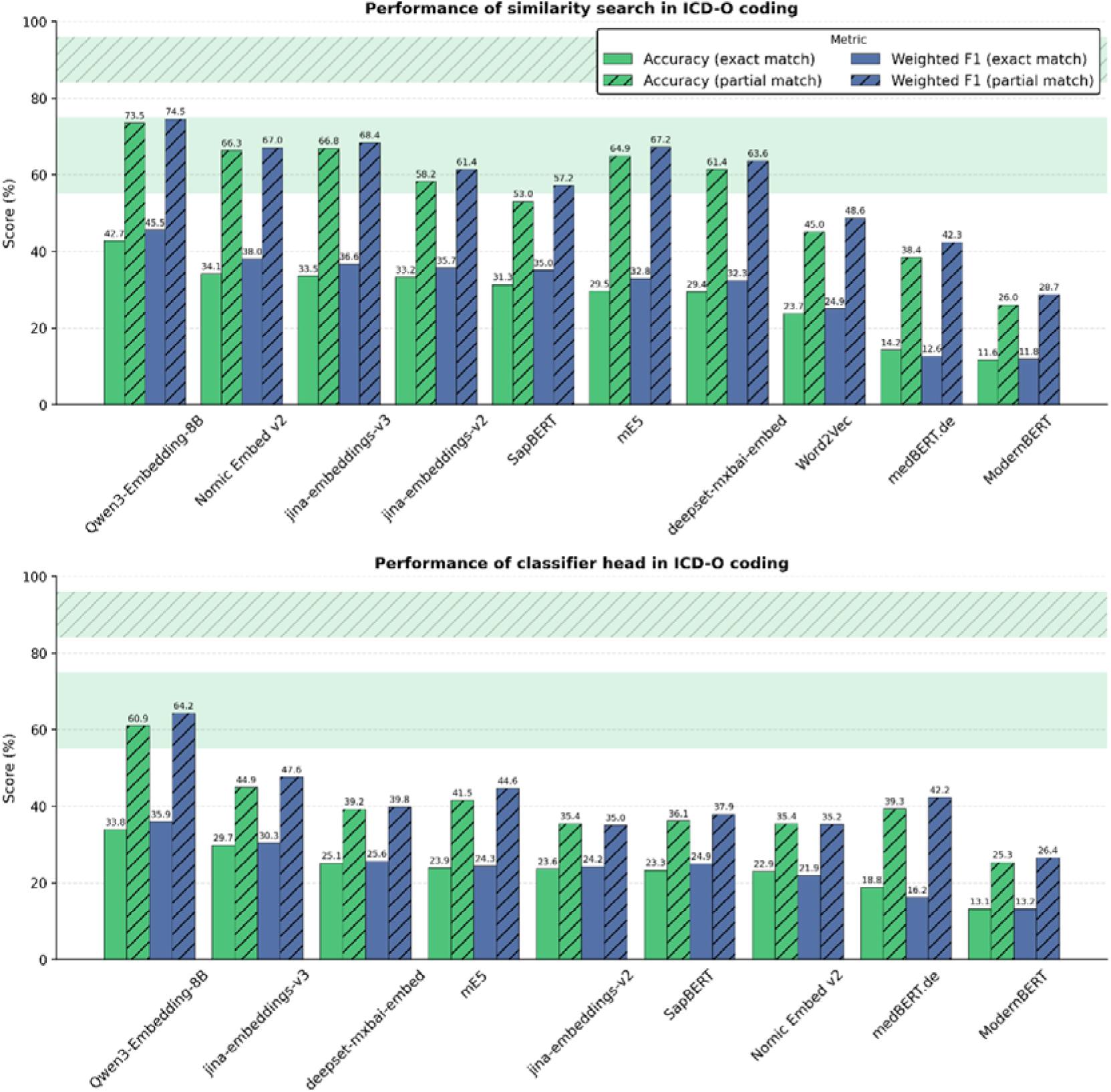
Comparison of similarity search (top) and classification (bottom) using embedding models for ICD-O coding. The performance on the test set is evaluated based on accuracy and weighted F1 for exact match and partial match. The models are ordered by decreasing exact match accuracy. The green bands show the 95% confidence interval for the maximum attainable accuracy according to the data quality analysis. The upper dashed band indicates the partial match accuracy limit and the lower band the exact match accuracy limit.

In accordance with the ICD-10 findings, the partial-match performance for ICD-O coding exceeded the exact-match performance by about 25–35 percentage points across all models in similarity search, indicating that errors are again predominantly driven by fine-grained code differentiation rather than complete semantic mismatch.

Furthermore, the Word2Vec baseline displayed low performance in both coding tasks. However, it did not rank last for ICD-O coding and outperformed the domain-specific medBERT.de model, thereby underscoring the notion that domain pretraining alone does not guarantee superior retrieval performance.

Across all the embedding models, training a classifier led to an approximate average gain of 4.7% for the exact match and a gain of 2% for the partial match, compared to the similarity search. For ICD-O coding, the reverse effect was observed. There, the classifier performed worse (approximately 5% for exact match and 16.7% for partial match accuracy).

From a practical standpoint, the classifier-based approach is more computationally efficient during inference. Predictions with the classifier were made in an average of 0.02 ms per sample (0.44 ms including embedding), whereas ChromaDB-based retrieval took an average of 0.30 ms (1.16 ms including embedding).

The performance results of all embedding models with all possible combinations are available in Table S6 in the Supplemental Material.

### Classifier-head confidence profiles across prediction ranks

The number of examples retrieved and incorporated into the final prompt for the LLM was determined by the classifier head’s overall distribution of the confidence scores. The confidence scores are computed as softmax probabilities at the output layer of the neural network classifier built on top of the embedding model. They represent the probabilities assigned to the predicted classes. Figure 5 summarizes the distribution of these probabilities across the top ten predictions. The probability distribution for both ICD-10 and ICD-O is observed to be sharply concentrated at rank 1, with a rapid decay in intensity as the rank increases. This finding suggests that the model predominantly allocates the majority of its probability mass to the top prediction, while assigning a comparatively smaller amount to lower-ranked alternatives. ICD-O demonstrates a broader rank-1 distribution and more substantial probability mass at rank 2 compared with ICD-10, reflecting a higher degree of ambiguity among the most plausible candidates in the topography setting. The rapid decay in confidence after the first few ranks suggested that only a small number of candidates need to be included in the RAG prompt. We therefore chose to include the top three retrieved codes, as these captured the most plausible alternatives.

**Figure 5.**
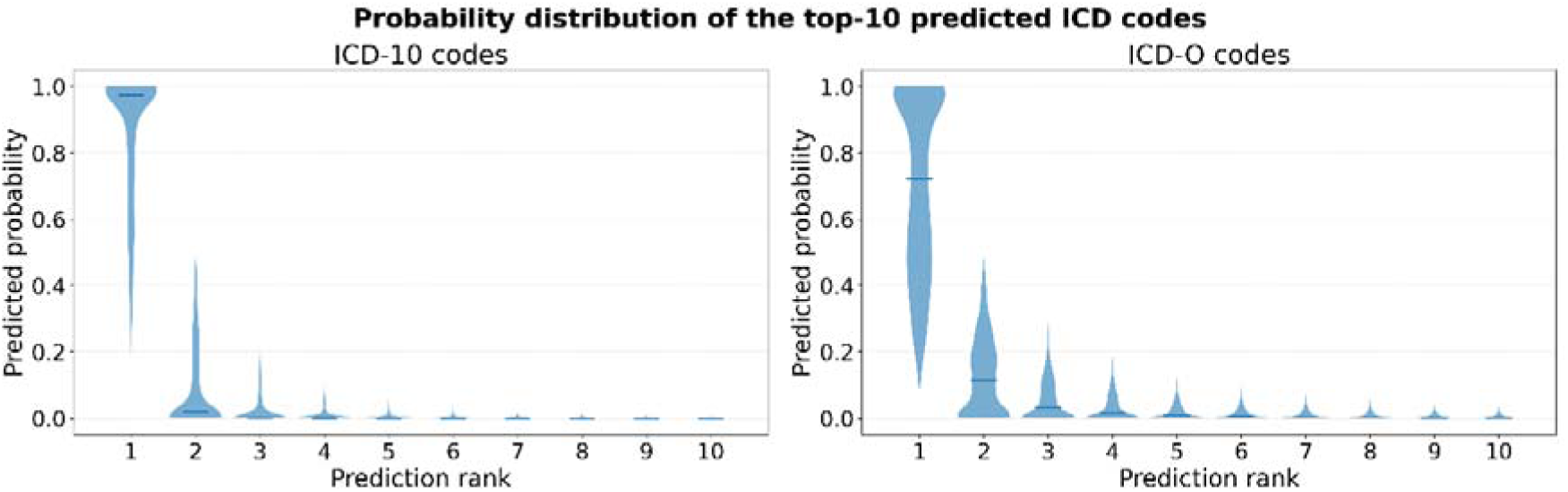
Violin plots of the predicted probability distributions produced by the trained classifier head for the top-10 ranked outputs in ICD-10 (left) and ICD-O (right) coding, illustrating the concentration of probability mass across prediction ranks.

### Comparison of RAG performance with embedding approaches and fine-tuned LLMs

We next evaluated the performance of base and fine-tuned Llama 3.1 8B and Llama 3.3 70B models across different prompting settings to assess the effects of model size, fine-tuning, and retrieval support on coding accuracy. For the retrieval component of the RAG pipeline, the Qwen3-Embedding-8B embedding model was selected based on its superior performance in the preceding retrieval experiments. Figure 6 summarizes the LLM performance across different prompting strategies and compares these results with the previously described retrieval baselines. Here, Llama 3.3 70B consistently outperformed Llama 3.1 8B across all prompting settings. Similar to the comparison of embedding models, the bigger model yielded the best results.

**Figure 6.**
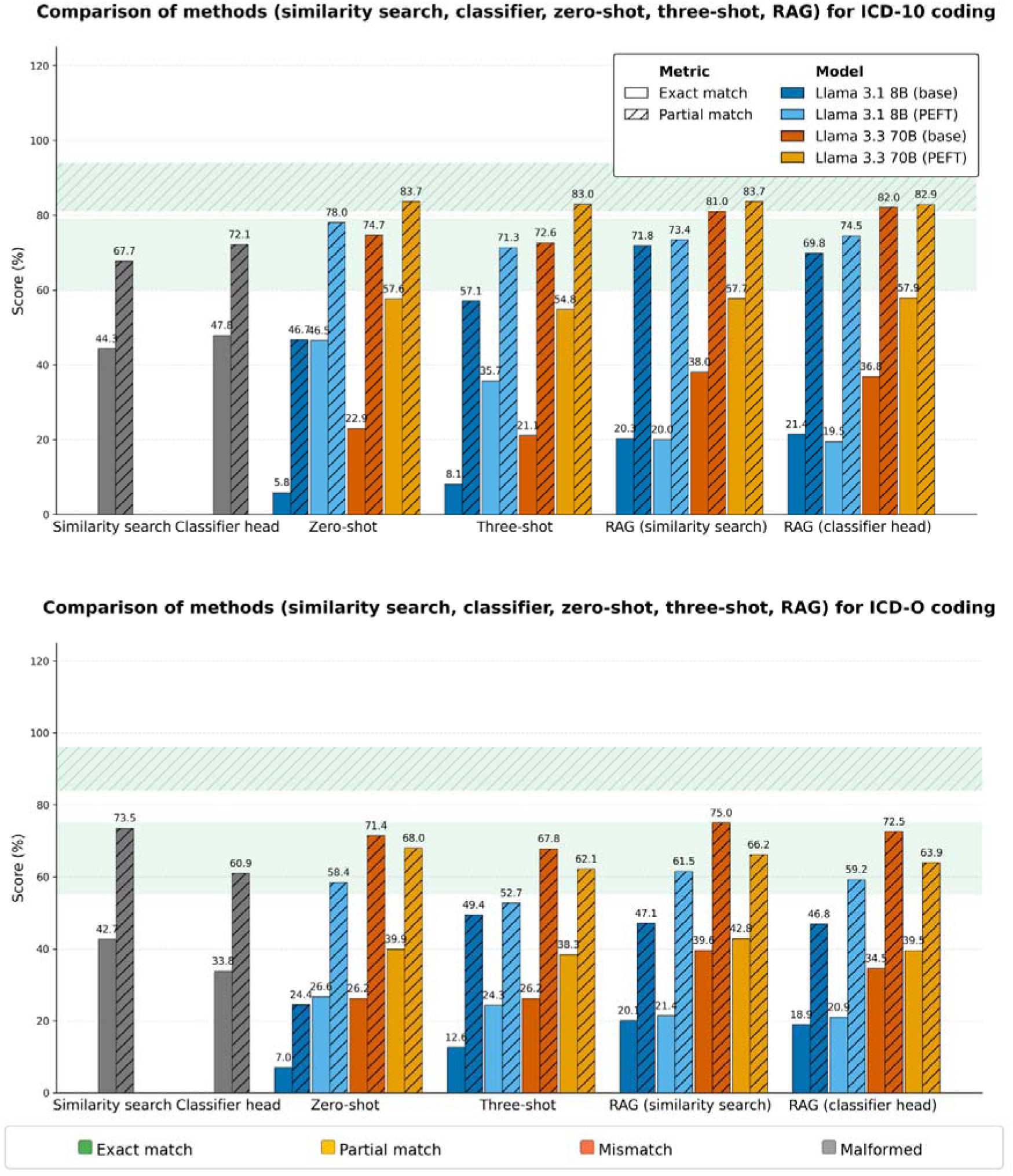
Comparison of LLM performance for ICD-10 (top) and ICD-O coding (bottom). Bar plots show exact- and partial-match accuracy for similarity search and classifier head baselines, as well as zero-shot, few-shot, and retrieval-augmented generation (RAG) prompting. Results are reported for Llama 3.1 8B (base and PEFT) and Llama 3.3 70B (base and PEFT).

Model size had a pronounced impact on automatic ICD code assignment in our setting. Llama 3.3 70B substantially outperformed Llama 3.1 8B, with the best-performing base configuration attaining 38.0% exact-match accuracy for ICD-10 and 39.6% for ICD-O with RAG prompting. However, none of the configurations reached the 95% confidence interval for exact match accuracy for either ICD-10 or ICD-O.

Notably, the base versions of the LLMs did not surpass the pure embedding-based approaches in terms of exact-match performance for either ICD-10 or ICD-O. Without any in-context examples, the performance of the base models was very low. Considering only the base LLMs, only Llama 3.3 70B could improve upon the partial-match performance of the embedding models for ICD-10 coding. The improvement was present both in zero-shot prompting and with RAG prompting.

For ICD-O, analogous gains were observed across prompting techniques, while the overall performance was lower than for ICD-10. In this context, base Llama 3.3 70B augmented with RAG outperformed the retrieval-only similarity-search baseline, achieving 75% partial-match accuracy. This result exceeded the retrieval baseline by 1.5 percentage points and even exceeded the fine-tuned model on this metric.

For ICD-10 coding, the performance achieved with RAG prompting was not sufficient to surpass the fine-tuned variants under exact-match evaluation. Regarding partial match, however, RAG closed much of the gap and approached the fine-tuned performance. Notably, Llama 3.3 70B even reached the reported confidence interval for ICD-10 partial code derivation. The fine-tuned models performed better with an exact-match accuracy of 42.8%, thereby surpassing the retrieval baseline.

Although the embedding models exhibited superior performance in comparison to base LLMs, the employment of RAG prompting led to a substantial enhancement in the performance of the base LLMs. This enhancement resulted in an increase of approximately 13 percentage points in exact-match accuracy and 11 percentage points in partial-match accuracy for ICD-10 coding. Base Llama 3.1 8B demonstrated the largest partial-match gains with RAG, highlighting the significance of external evidence for smaller base models. In contrast, the improvement for Llama 3.3 70B was more modest, yet still sufficient to increase ICD-10 partial-match performance to 80.6% and meet the predefined confidence-band criterion. For ICD-O coding and averaged across all embedding models, RAG prompting led to an increase of approximately 6-8 percentage points in exact-match accuracy and 6 percentage points in partial-match accuracy. In this setting, the base Llama 3.3 70B model achieved the best result across all model configurations with RAG based on similarity search.

For the base LLMs, three-shot prompting generally outperformed zero-shot prompting. The comparison of three-shot prompting with random examples to the three-shot prompting with retrieval-informed examples in RAG indicates that the gains are not attributable to the prompt format alone, but that the models draw information about possible codes from the examples. A notable exception was the fine-tuned Llama 3.1 8B in exact-match evaluation, where the exact-match accuracy declined from three-shot to RAG prompting while the partial-match performance remained comparatively more robust. In contrast, the base Llama 3.1 8B improved under RAG.

In the ICD-10 setting, fine-tuned models achieved the strongest performance under partial-match evaluation and consistently outperformed both embedding-only approaches. Under exact-match evaluation, however, this advantage was restricted to the fine-tuned Llama 3.3 70B, which also outperformed both similarity search and the classifier head. In the ICD-O setting, Llama 3.1 8B followed the same pattern as in ICD-10, whereas the base Llama 3.3 70B surpassed its fine-tuned counterpart across all three prompting settings regarding the partial-match accuracy.

To further assess class-wise performance, we compared macro F1 and weighted F1 scores. The corresponding results are reported in Tables A5 and A6 of the Supplemental Material, for ICD-10 and ICD-O-3 respectively. Across all models and for both coding tasks, weighted F1 consistently exceeded macro F1 by a substantial margin. This pattern is consistent with class imbalance and suggests that the models performed better on frequent codes than on rare ones. The largest gaps were observed for Llama 3.3 70B PEFT, with a difference of approximately 22 percentage points in the Qwen3-Embedding-8B setting, indicating particularly strong performance concentration on prevalent diagnoses. A similar pattern was observed for the similarity-search baselines, which showed gaps of around 20 percentage points across both coding schemes. In contrast, Llama 3.1 8B PEFT exhibited smaller differences between weighted and macro F1. However, this narrower gap appears to reflect generally weak performance across both frequent and infrequent codes rather than more balanced class-wise predictions.

Although retrieval augmentation improved overall performance, particularly for base Llama 3.3 70B, the gains were larger for weighted F1 than for macro F1. This suggests that retrieval primarily improved performance on more frequent classes. Compared with ICD-10, ICD-O-3 generally yielded lower macro F1 scores; for example, with RAG and Qwen3-Embedding-8B, macro F1 reached 40.87% for ICD-10 but only 30.54% for ICD-O-3 in the Llama 3.3 70B PEFT setting.

In terms of execution time, per-sample inference for answering the prompts remained below one second per prompt, but was still approximately 1000 times slower than the pure retrieval step (see Table S5 of the Supplemental Material for all LLM inference times).

### Impact of retrieval quality on final LLM-assisted ICD prediction

To assess the influence of retrieval quality on RAG output, we traced the quality of retrieved codes and compared it with the final LLM predictions in Sankey plots (Figure 7). For both ICD-10 and ICD-O coding, the codes retrieved from the embedding models are mostly correct (62.6% / 58.6% exact matches in top-3 predictions of ICD-10 / ICD-O codes, and only 15.9% / 15.4% without any code suggestions that match at least partly). Surprisingly, the LLMs could not translate these suggestions into equally good predictions and pick the correct code from the options presented to them. A large portion of code suggestions with exact matches were finally answered with a code that matches only partly. From the Sankey plots, it also becomes clearer that base Llama 3.1 8B exhibited a large portion of malformed outputs in the ICD-O coding task. In all other cases, the rate of malformed outputs was very low and therefore negligible.

**Figure 7.**
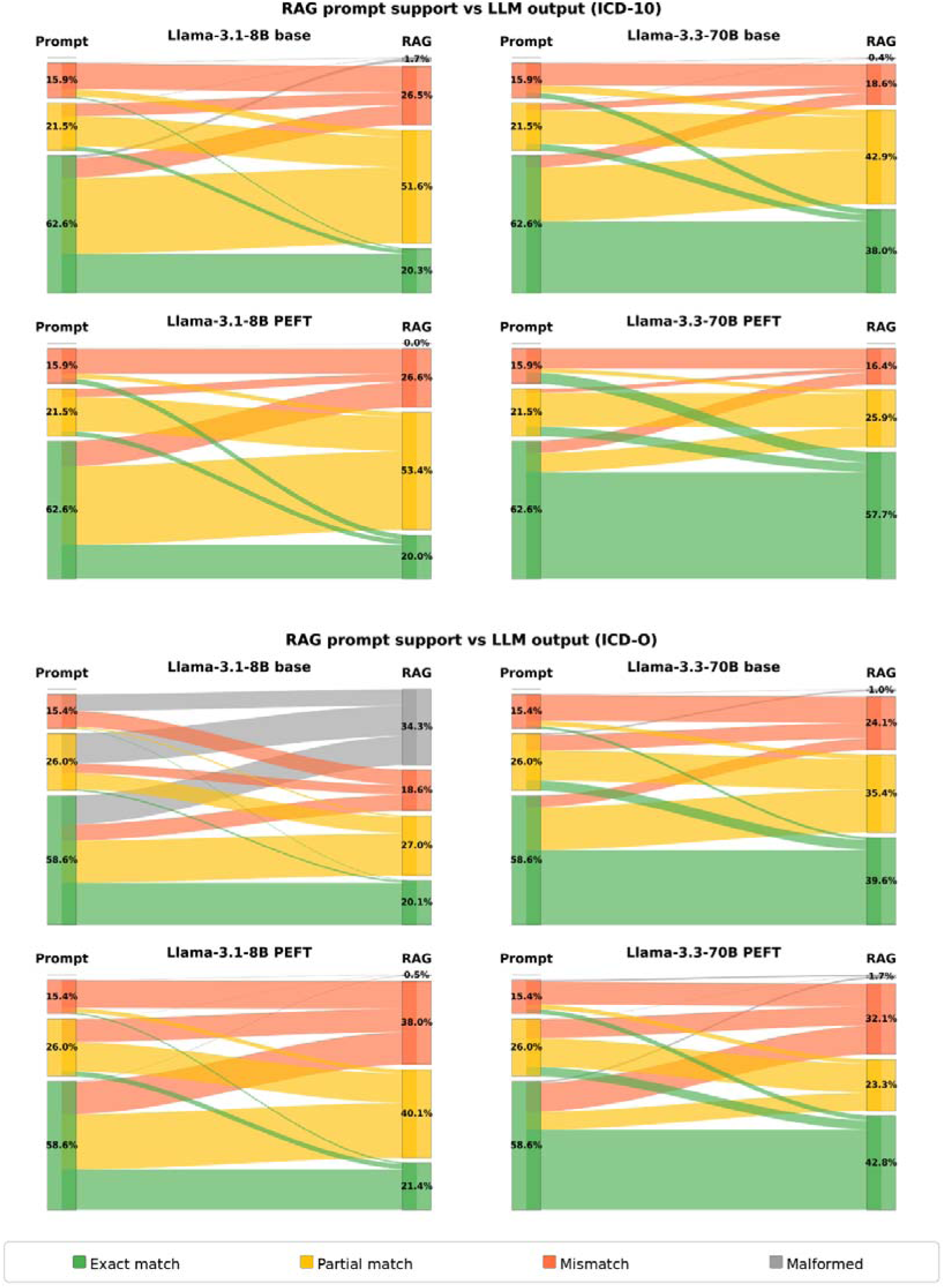
Sankey plots illustrate how retrieval support influences prediction outcomes for ICD-10 (top) and ICD-O-3 (bottom) coding. The left columns in the Sankey plots indicate whether the retrieved codes contain the exact code, at least a partially matching code, or no matching code at all. The right columns show whether the final model answer was an exact match, partial match, mismatch, or could not be parsed (malformed). Flows represent transitions from prompt-level support categories to final model outputs. Results are shown for Llama 3.1 8B and Llama 3.3 70B, in base and PEFT configurations.

## Discussion

Our study systematically compares key methodological components relevant to RAG-based ICD coding, focusing on a diverse set of embedding architectures and two retrieval strategies, examined both independently and in combination with RAG. Accordingly, we evaluated nine embedding models, and Word2Vec as an additional baseline, and selected two LLMs from the Llama family on the basis of prior work ^5^. This design enabled a focused analysis of retrieval and prompting effects without introducing a large number of model combinations in the RAG setting. Given the rapid evolution of the LLM landscape, we considered this targeted comparison more informative than a broad benchmark across many models that might quickly become outdated. Nevertheless, restricting the analysis to a single LLM family remains a limitation, and future work should examine in greater detail how model family, training regime, and model size influence RAG performance in this setting.

Coding German diagnosis descriptions with RAG and embedding models has been explored by other studies as well. Kreuzthaler et al. ^4^ employed SapBERT-UMLS and medBERT.de on a knowledge base comprising 16.4 million diagnostic descriptions from an Austrian hospital network. In their evaluation, they achieved weighted F1 scores of 0.86 and 0.87 for an exact match of the ICD-10 code via embedding-based nearest neighbor classification. Using only standardized terminology from the Austrian ICD-10 BMSGPK dataset, similar to our approach, they achieved weighted F1 scores of 0.61 and 0.55, respectively. In our study, medBERT.de achieved a weighted F1 score of 0.30, whereas Qwen3-Embedding-8B achieved 0.49. Coding performance might be further improved by incorporating real-world medical data into the knowledge base, rather than relying exclusively on public classification data. However, the primary objective of our study was to undertake a comparative analysis of methodologies.

Krumscheid et al. ^6^ employed a multi-stage, retrieval-augmented generation pipeline to analyze unstructured German cardiology letters. They achieved a top-1 accuracy of 76.3% for an exact match and 78.1% for partial matches across 228 diagnoses and 122 ICD-10 codes. Although the RAG pipeline for ICD-10 coding was conceptually similar, the codes in the study were obtained by sampling ten German doctors’ letters, which represents a small and manually annotated set.

Böhringer et al. ^19^ used a Word2Vec embedding model for ICD-10 coding of ophthalmological doctors’ letters, achieving 69% exact-match accuracy and 86%-91% partial-match accuracy when diagnosis descriptions from one hospital were used as the knowledge base and the model was evaluated on data from two other hospitals. Consistent with prior work, simpler embedding-based retrieval approaches can outperform LLMs in some medical coding settings. In our study, Word2Vec outperformed Llama 3.1 8B, although the larger Llama 3.3 70B surpassed the Word2Vec baseline. Nevertheless, Word2Vec achieved only 18.8% exact-match accuracy and 39.0% partial-match accuracy and was also outperformed by the larger embedding models included in our study. One possible explanation for the discrepancy compared with the findings of Böhringer et al. is that coding tumor diagnoses may be more challenging than coding ophthalmological diagnoses.

Our study was focused on a clear comparison of embedding methods and RAG approaches rather than maximizing the performance for a very specific use case. When comparing the two pure embedding-based approaches, semantic search and classifier head, the classifier achieved better results for ICD-10 coding but the semantic search was better in ICD-O coding. This may be due to less available data (approximately 25% less) and also due to the construction of the diagnosis descriptions in the knowledge base, which were constructed from the Alpha-ID diagnosis descriptions and the location appended in parentheses. This may lead to closer embeddings that are more difficult to separate with ReLU layers. In such dense clusters, neural networks can struggle to define robust decision boundaries, especially with sparse data^41^. Since sentence embeddings prioritize semantic proximity over linear separability ^17^, non-parametric semantic search remains more robust to fine-grained overlaps by bypassing the need for explicit boundary learning. While the results of the standalone methods differed for ICD-10 and ICD-O coding, these differences became far less pronounced when the methods were incorporated into the RAG pipeline.

To further compare the methods, we included fine-tuned model variants to assess the impact of fine-tuning on RAG performance and to compare the RAG performance directly with these adapted models. Interestingly, while the fine-tuned version of Llama 3.3 70B achieved the best performance for ICD-10 coding, the base model performed best in the ICD-O setting. One reason for this can probably be found in the training dataset used for fine-tuning, which had a lower variety of ICD-O diagnosis descriptions than what was used in the present study ^5^.

Alongside our code, we also publish the dataset that we created from a combination of the Alpha-ID classification and translation tables to ICD-O issued by a consortium of German cancer registries. This dataset served here as the basis for semantic similarity search and classification for ICD-O coding. Since we relied on publicly available data, the dataset can be shared freely, allowing the reproducibility of our methods. Thus, the dataset can also be reused on its own as a knowledge base for ICD-O topography coding in other projects.

Strengthening the data basis for automated ICD-O coding is especially important, as we attribute the comparatively lower performance observed for ICD-O coding to model- and data-related factors. Differences in pre-training exposure likely create a large portion of this gap, with information about the ICD-10 being more abundant than knowledge about the ICD-O classification, which is more specialized and less represented in typical pre-training corpora ^5^. This limited prior knowledge may also be a reason for the frequent confusion between localization (ICD-O topography) and histological information (ICD-O morphology) observed in model answers, resulting in a relevant proportion of hallucinations and formatting errors in ICD-O predictions. Future work could also further integrate the full semantic potential of the ICD-O classification by including the morphology coding. For this purpose, however, more details about the histopathology are often required than given in the short free-text diagnosis descriptions used for this study. One further relevant consideration is that the knowledge base was derived from standardized and public classification datasets, whereas evaluation was conducted on real-world clinical data. This design supports the assessment of generalizability across different hospitals, although differences in terminology, label consistency, and documentation complexity between public and routine clinical data may still introduce representation-related bias.

One additional limitation of our study is that we did not systematically evaluate different prompt phrasings or prompt designs. Instead, we used the same prompting questions as in the fine-tuned variants. This facilitated the comparison between base and fine-tuned models, as the performance of fine-tuned models may degrade if the prompt format differs ^42^. We did, however, vary the information basis and compare model performance across zero-shot, three-shot, and retrieval-informed RAG prompts. In few-shot prompting, we accounted for recency bias by ordering the suggestions with ascending probability ^43^. We could verify this bias existed as changing to this order resulted in a small gain in accuracy.

While RAG improved LLM performance in most settings, correct retrieved codes were often still not selected in the final output. This suggests that the main challenge of RAG in this setting is not retrieval quality alone, but the model’s ability to incorporate retrieved suggestions reliably into the final coding decision. This challenge was particularly evident for the smaller Llama 3.1 8B model, which was prone to generate malformed or hallucinated outputs. Investigating different prompting techniques to increase the integration of retrieved knowledge could be a promising direction. Additionally, fine-tuning LLMs for better integrating the retrieved knowledge could be another way to improve the performance ^5^. Training models directly on structured RAG-prompts may improve their ability to integrate retrieved information, as suggested in recent studies on supervised RAG and fine-tuned LLMs ^44^. Such approaches could further enhance the combined performance of retrieval-based and fine-tuning strategies for complex clinical coding problems.

Hou et al. identified abbreviations and related lexical variations as major error sources in real-world clinical text and explicitly addressed them as a key robustness challenge for LLM-based coding ^45^. By expanding abbreviations in our inputs, we minimized confounding factors that can disproportionately harm vanilla models and thereby improved the interpretability of performance differences attributable to the coding task itself rather than surface-form noise ^45^.

## Conclusion

In conclusion, we evaluated automated ICD-10 and ICD-O-3 topography coding for German tumor diagnoses using different embedding models and retrieval-augmented generation (RAG) with Llama 3.1 8B and Llama 3.3 70B, and corresponding fine-tuned variants.

Our results show that RAG prompting substantially improved the performance of base LLMs. To fully leverage the RAG approach, both a strong embedding model and a strong LLM need to be selected. While RAG provided advantages over a pure embedding-based approach in some settings, such as higher partial ICD-10 code match accuracy, it did not consistently outperform the embedding-only approach across all metrics. Particularly, the exact match accuracy with RAG did not exceed the embedding-based approaches. Given that RAG is more resource-intensive and introduces additional system complexity, its use should be evaluated carefully in the specific application setting. In this context, a direct comparison with embedding-only approaches, as performed here, is essential to determine whether the added effort translates into meaningful performance gains.

Additionally, our results also show differences between embedding models. Across the embedding models, stronger performance was observed for larger general-purpose models such as Qwen3-Embedding-8B than for smaller medically adapted models such as medBERT.de and SapBERT, suggesting that model size may be more important than medical-domain adaptation. A promising direction for future work could be to fine-tune such embedding models on medical datasets to further improve their task-specific performance and reach performances that are practically usable. Fine-tuning LLMs to make better use of retrieved information could be a valuable next step, given that the models did not consistently incorporate the suggestions generated by the embedding models.

## Authors’ contributions

F.A. conducted the experiments and wrote the initial draft of the manuscript. S.L. conceived the study, directed the experimental design and revised the manuscript. A.U. conducted experiments that were integrated in the evaluation. L.O.R contributed to the development of the evaluation software. T.K. provided the data from the tumor documentation system, and G.V. performed the data extraction. T.P. provided the funding and supervised the work. All authors read and approved the final version of the manuscript.

## Statements and Declarations

### Ethical considerations

The requirement for an ethics approval was waived by the responsible ethics commission on 06 September 2024 because only short diagnosis descriptions and codes without identifying information were used for this study.

### Consent to participate

Not applicable

### Consent for publication

Not applicable

### Declaration of conflicting interest

The authors declared no potential conflicts of interest with respect to the research, authorship, and/or publication of this article.

### Funding statement

This research has been supported by the Federal Ministry of Research, Technology and Space (BMFTR) in Germany in the project “Digitaler FortschrittsHub Gesundheit – DECIDE” (FKZ 01ZZ2106A) of the German national medical informatics initiative. The project aims to facilitate data transfer and interoperability among different stakeholders in the healthcare sector and to provide decision support based on structured patient information.

### Data availability statement

The code, training datasets, and results are available at https://github.com/Fatma-Alic/tumor-icd-coding-rag.

## Notes

### Competing Interest Statement

The authors have declared no competing interest.

### Author Declarations

The Ethics Committee of the State Medical Association of Rhineland-Palatinate waived ethical approval for this work.

## References

1. Bekanntmachung - Aktualisierter einheitlicher onkologischer Basisdatensatz der Arbeitsgemeinschaft Deutscher Tumorzentren e. V. (ADT) und der Gesellschaft der epidemiologischen Krebsregister in Deutschland e. V. (GEKID).

2. Federal Institute for Drugs and Medical Devices (BfArM). ICD-10-GM Alphabetical index. 2024. Accessed March 7, 2025. https://www.bfarm.de/EN/Code-systems/Classifications/ICD/ICD-10-GM/Alphabetical-index/_node.html

3. Federal Institute for Drugs and Medical Devices (BfArM). ICD-O-3: International Classification of Diseases for Oncology. 2024. Accessed March 7, 2025. https://www.bfarm.de/EN/Code-systems/Classifications/ICD/ICD-O-3/_node.html

4. Kreuzthaler M, Pfeifer B, Schulz S. Secondary Use of Clinical Problem List Descriptions for Bi-Encoder Based ICD-10 Classification. https://pubmed.ncbi.nlm.nih.gov/40417589/

5. Lenz S, Rosario LO, Vollmar G, et al. Unlocking Public Catalogues: Instruction-Tuning LLMs for ICD Coding of German Tumor Diagnoses. *arXiv*. Preprint posted online October 15, 2025:arXiv:2510.13624. doi:10.48550/arXiv.2510.13624

6. Krumscheid M, Blömer J, Becker M. Retrieval-Augmented Generation for ICD-10 Coding in German Clinical Texts – A Technical Case Report. In: Röhrig R, Ganslandt T, Jung K, et al., eds. Studies in Health Technology and Informatics. IOS Press; 2025. doi:10.3233/SHTI251397

7. Devlin J, Chang MW, Lee K, Toutanova K. BERT: Pre-training of Deep Bidirectional Transformers for Language Understanding. arXiv. Preprint posted online May 24, 2019:arXiv:1810.04805. doi:10.48550/arXiv.1810.04805

8. Bressem KK, Papaioannou JM, Grundmann P, et al. medBERT.de: A comprehensive German BERT model for the medical domain. Expert Syst Appl. 2024;237:121598. doi:10.1016/j.eswa.2023.121598

9. Lenz S, Ustjanzew A, Jeray M, Ressing M, Panholzer T. Can open source large language models be used for tumor documentation in Germany?—An evaluation on urological doctors’ notes. BioData Min. 2025;18(1):48. doi:10.1186/s13040-025-00463-8

10. Dubey A, Jauhri A, Pandey A, et al. The Llama 3 Herd of Models. *arXiv*. Preprint posted online July 31, 2024:arXiv:2407.21783. Accessed August 1, 2024. http://arxiv.org/abs/2407.21783

11. Gallifant J, Afshar M, Ameen S, et al. The TRIPOD-LLM reporting guideline for studies using large language models. Nat Med. 2025;31(1):60–69. doi:10.1038/s41591-024-03425-5

12. Federal Institute for Drugs and Medical Devices (BfArM). Alpha-ID-SE Version 2024 EDV-Fassung TXT (CSV). Accessed November 12, 2025. https://www.bfarm.de/SharedDocs/Downloads/DE/Kodiersysteme/klassifikationen/alpha-id/version2024/alphaidse2024_zip.html?nn=841246&cms_dlConfirm=true&cms_calledFromDoc=841246

13. BfArM - ICD-10-GM. Accessed November 12, 2025. https://www.bfarm.de/EN/Code-systems/Classifications/ICD/ICD-10-GM/_node.html

14. Umsetzungsleitfaden - Umsetzungsleitfaden - Plattform § 65c - Confluence Instanz. Accessed November 26, 2025. https://plattform65c.atlassian.net/wiki/spaces/UMK/overview?homepageId=15532036

15. Blaming clinical-abbreviations/metainventory/Metainventory_Version1.0.0.csv at master · lisavirginia/clinical-abbreviations. GitHub. Accessed November 12, 2025. https://github.com/lisavirginia/clinical-abbreviations

16. Mohr I, Krimmel M, Sturua S, Akram MK, Koukounas A. Model card of jina-embeddings-v2-base-de on Hugging Face. November 1, 2025. Accessed January 13, 2026. https://huggingface.co/jinaai/jina-embeddings-v2-base-de

17. Reimers N, Gurevych I. Sentence-BERT: Sentence Embeddings using Siamese BERT-Networks. In: Proceedings of the 2019 Conference on Empirical Methods in Natural Language Processing. Association for Computational Linguistics; 2019. https://arxiv.org/abs/1908.10084

18. Harbecke D, Chen Y, Hennig L, Alt C. Why only Micro-F1? Class Weighting of Measures for Relation Classification. In: Shavrina T, Mikhailov V, Malykh V, Artemova E, Serikov O, Protasov V, eds. Proceedings of NLP Power! The First Workshop on Efficient Benchmarking in NLP. Association for Computational Linguistics; 2022:32–41. doi:10.18653/v1/2022.nlppower-1.4

19. Böhringer D, Angelova P, Fuhrmann L, et al. Automatic inference of ICD-10 codes from German ophthalmologic physicians’ letters using natural language processing. Sci Rep. 2024;14(1):9035. doi:10.1038/s41598-024-59926-3

20. chromadb: Chroma. Accessed March 23, 2026. https://github.com/chroma-core/chroma

21. Schmidt B. bmschmidt/wordVectors. Published online March 13, 2017. Accessed March 4, 2026. https://github.com/bmschmidt/wordVectors

22. Keil JM. Efficient Bounded Jaro-Winkler Similarity Based Search. Published online 2019. doi:10.18420/BTW2019-13

23. Muennighoff N, Tazi N, Magne L, Reimers N. MTEB: Massive Text Embedding Benchmark. *arXiv*. Preprint posted online March 19, 2023:arXiv:2210.07316. doi:10.48550/arXiv.2210.07316

24. Zhang Y, Li M, Long D, et al. Model card of Qwen3-Embedding-8B on Hugging Face. November 1, 2025. Accessed October 27, 2025. https://huggingface.co/Qwen/Qwen3-Embedding-8B

25. Sturua S, Mohr I, Akram MK, et al. Model card of jina-embeddings-v3 on Hugging Face. November 1, 2025. Accessed October 27, 2025. https://huggingface.co/jinaai/jina-embeddings-v3

26. Wang L, Yang N, Huang X, Yang L, Majumder R, Wei F. Model card of mE5 on Hugging Face. November 1, 2025. Accessed October 27, 2025. https://huggingface.co/intfloat/multilingual-e5-large-instruct

27. Lee S, Shakir A, König D, Lipp J. Model card of deepset-mxbai-embed on Hugging Face. November 1, 2025. Accessed October 27, 2025. https://huggingface.co/mixedbread-ai/deepset-mxbai-embed-de-large-v1

28. Nussbaum Z, Duderstadt B. Model card of Nomic-Embed-v2 on Hugging Face. November 1, 2025. Accessed October 27, 2025. https://huggingface.co/nomic-ai/nomic-embed-text-v2-moe

29. Mustafa FE, Dima C, Ochoa J, Staab S. Model card of SapBERT on Hugging Face. November 1, 2025. Accessed October 27, 2025. https://huggingface.co/permediq/SapBERT-DE

30. Remy F. Model card of ModernBERT on Hugging Face. November 1, 2025. Accessed October 27, 2025. https://huggingface.co/Parallia/Fairly-Multilingual-ModernBERT-Embed-BE-DE

31. Bressem KK, Papaioannou JM, Grundmann P, et al. Model card of medMERT.de on Hugging Face. November 1, 2025. Accessed October 27, 2025. https://huggingface.co/GerMedBERT/medbert-512

32. Cui Y, Jia M, Lin TY, Song Y, Belongie S. Class-Balanced Loss Based on Effective Number of Samples. https://arxiv.org/abs/1901.05555

33. Falis M, Pajak M, Lisowska A, et al. Ontological attention ensembles for capturing semantic concepts in ICD code prediction from clinical text. In: Holderness E, Jimeno Yepes A, Lavelli A, Minard AL, Pustejovsky J, Rinaldi F, eds. Proceedings of the Tenth International Workshop on Health Text Mining and Information Analysis (LOUHI 2019). Association for Computational Linguistics; 2019:168–177. doi:10.18653/v1/D19-6220

34. Loshchilov I, Hutter F. Decoupled Weight Decay Regularization. In: 7th International Conference on Learning Representations, ICLR 2019, New Orleans, LA, USA, May 6-9, 2019. OpenReview.net; 2019. https://openreview.net/forum?id=Bkg6RiCqY7

35. Meta, Inc. Model card of Llama 3.1 8B on Hugging Face. December 6, 2024. Accessed January 15, 2025. https://huggingface.co/meta-llama/Llama-3.1-8B-Instruct

36. Lenz S, Rosario LO. Model card of Llama-3.1-8B-ICDOPS-QA-2024 on Hugging Face. October 16, 2025. Accessed April 20, 2026. https://huggingface.co/stefan-m-lenz/Llama-3.1-8B-ICDOPS-QA-2024

37. Meta AI. Model card of Llama 3.3 70B on Hugging Face. December 6, 2024. Accessed August 19, 2025. https://huggingface.co/meta-llama/Llama-3.3-70B-Instruct

38. Lenz S, Rosario LO. Model card of Llama-3.3-70B-ICDOPS-QA-2024 on Hugging Face. October 16, 2025. Accessed April 20, 2026. https://huggingface.co/stefan-m-lenz/Llama-3.3-70B-ICDOPS-QA-2024

39. Houlsby N, Giurgiu A, Jastrzebski S, et al. Parameter-Efficient Transfer Learning for NLP. In: Proceedings of the 36th International Conference on Machine Learning. PMLR; 2019:2790–2799. Accessed April 17, 2026. https://proceedings.mlr.press/v97/houlsby19a.html

40. Hu EJ, Shen Y, Wallis P, et al. LoRA: Low-Rank Adaptation of Large Language Models. In: 2021. Accessed April 17, 2026. https://openreview.net/forum?id=nZeVKeeFYf9

41. Snell J, Swersky K, Zemel R. Prototypical Networks for Few-shot Learning. In: Advances in Neural Information Processing Systems. Vol 30. Curran Associates, Inc.; 2017. Accessed April 1, 2026. https://papers.nips.cc/paper_files/paper/2017/hash/cb8da6767461f2812ae4290eac7cbc42-Abstract.html

42. Yuan M, Shing HC, Strong M, Shivade C. Toward Reliable Clinical Coding with Language Models: Verification and Lightweight Adaptation. In: Potdar S, Rojas-Barahona L, Montella S, eds. Proceedings of the 2025 Conference on Empirical Methods in Natural Language Processing: Industry Track. Association for Computational Linguistics; 2025:173–184. doi:10.18653/v1/2025.emnlp-industry.12

43. Zhao TZ, Wallace E, Feng S, Klein D, Singh S. Calibrate Before Use: Improving Few-Shot Performance of Language Models. https://arxiv.org/abs/2102.09690

44. Pingua B, Sahoo A, Kandpal M, et al. Medical LLMs: Fine-Tuning vs. Retrieval-Augmented Generation. Bioengineering. 2025;12(7):687. doi:10.3390/bioengineering12070687

45. Hou Z, Liu H, Bian J, He X, Zhuang Y. Enhancing medical coding efficiency through domain-specific fine-tuned large language models. Npj Health Syst. 2025;2(1):14. doi:10.1038/s44401-025-00018-3

