## Supplemental Material 1 for "To RAG, or Not to RAG? A Comparative Evaluation of Retrieval-Augmented Generation for ICD Coding of German Tumor Diagnoses"

**Supplementary Material 1. Overview of embedding model results, training results, and prompt templates**

**Table S1** Training time of classifier head for ICD-10 coding. Each embedding model was trained for 70 epochs.

| Model | Training time [h] |
|---|---|
| Qwen3-Embedding-8B | 5.2 |
| jina-embeddings-v3 | 0.35 |
| mE5 | 0.36 |
| deepset-mxbai-embed | 0.36 |
| Nomic-Embed-v2 | 0.38 |
| SapBERT | 0.15 |
| jina-embeddings-v2 | 0.17 |
| ModernBERT | 0.22 |
| medBERT.de | 0.14 |

**Table S2** Training time of classifier head for ICD-O coding. Each embedding model was trained for 30 epochs.

| Model | Training time [h] | Learning rate | Batch size | Weight decay |
|---|---|---|---|---|
| Qwen3-Embedding-8B | 3.12 | $2.58 \times 10^{-4}$ | 32 | $4.28 \times 10^{-2}$ |
| jina-embeddings-v3 | 0.12 | $7.55 \times 10^{-5}$ | 16 | $7.30 \times 10^{-2}$ |
| mE5 | 0.16 | $4.03 \times 10^{-4}$ | 32 | $4.85 \times 10^{-3}$ |
| deepset-mxbai-embed | 0.17 | $1.43 \times 10^{-4}$ | 16 | $2.94 \times 10^{-2}$ |
| Nomic-Embed-v2 | 0.15 | $2.17 \times 10^{-4}$ | 16 | $3.44 \times 10^{-3}$ |
| SapBERT | 0.07 | $1.19 \times 10^{-4}$ | 16 | $5.55 \times 10^{-2}$ |
| jina-embeddings-v2 | 0.08 | $7.55 \times 10^{-5}$ | 16 | $7.30 \times 10^{-2}$ |
| ModernBERT | 0.07 | $3.43 \times 10^{-3}$ | 32 | $8.95 \times 10^{-2}$ |
| medBERT.de | 0.05 | $4.91 \times 10^{-4}$ | 16 | $2.54 \times 10^{-2}$ |

**Table S3** Results of Qwen3-Embedding-8B for ICD-10 coding for all prompting strategies and all LLM configurations.

| Method | LLM | Accuracy | Partial Accuracy | Weighted F1 (exact) | Weighted F1 (partial) | Macro F1 (exact) | Macro F1 (partial) |
|---|---|---|---|---|---|---|---|
| Sim. search | — | 44.27% | 67.74% | 48.70% | 70.72% | 30.27% | 49.74% |
| Classifer | — | 47.83% | 72.13% | 52.66% | 74.75% | 32.65% | 53.73% |
| Zero-shot | Llama 3.1 base | 5.58% | 44.47% | 4.56% | 45.97% | 3.72% | 25.13% |
| Zero-shot-prompting | Llama 3.1 PEFT | 45.50% | 78.11% | 50.53% | 79.71% | 29.10% | 55.74% |
| Zero-shot-prompting | Llama 3.3 base | 22.08% | 74.75% | 23.21% | 74.55% | 15.72% | 49.23% |
| Zero-shot-prompting | Llama 3.3 PEFT | 56.97% | 83.45% | 61.97% | 85.19% | 40.19% | 64.79% |
| Three-shot-prompting | Llama 3.1 base | 7.61% | 56.13% | 6.70% | 58.38% | 4.65% | 30.65% |
| Three-shot-prompting | Llama 3.1 PEFT | 35.18% | 71.54% | 40.13% | 72.40% | 21.16% | 47.51% |
| Three-shot-prompting | Llama 3.3 base | 20.75% | 72.13% | 22.24% | 73.47% | 16.26% | 50.60% |
| Three-shot-prompting | Llama 3.3 PEFT | 54.40% | 82.91% | 59.70% | 84.01% | 36.81% | 62.76% |
| RAG-prompting (similarity search) | Llama 3.1 base | 18.77% | 69.91% | 21.01% | 72.68% | 19.10% | 47.65% |
| RAG-prompting (similarity search) | Llama 3.1 PEFT | 20.70% | 73.86% | 25.34% | 75.47% | 18.66% | 48.85% |
| RAG-prompting (similarity search) | Llama 3.3 base | 38.04% | 80.88% | 41.77% | 82.20% | 27.70% | 60.81% |
| RAG-prompting (similarity search) | Llama 3.3 PEFT | 57.41% | 83.45% | 62.65% | 84.71% | 40.87% | 62.29% |
| RAG-prompting (classifier head) | Llama 3.1 base | 18.97% | 67.59% | 21.17% | 71.10% | 18.24% | 48.46% |
| RAG-prompting (classifier head) | Llama 3.1 PEFT | 20.16% | 73.91% | 24.04% | 75.74% | 19.70% | 49.34% |

| | | | | | | | |
|---|---|---|---|---|---|---|---|
| RAG-prompting (classifier head) | Llama 3.3 base | 36.76% | 81.42% | 40.92% | 83.11% | 25.87% | 62.09% |
| RAG-prompting (classifier head) | Llama 3.3 PEFT | 56.77% | 82.71% | 62.07% | 83.84% | 39.73% | 61.22% |

**Table S4** Results of Qwen3-Embedding-8B for ICD-O coding for all prompting strategies and all LLM configurations.

| **Method** | **LLM** | **Accuracy** | **Partial Accuracy** | **Weighted F1 (exact)** | **Weighted F1 (partial)** | **Macro F1 (exact)** | **Macro F1 (partial)** |
|---|---|---|---|---|---|---|---|
| Retrieval-baseline: similarity search | — | 42.69% | 73.47% | 45.46% | 74.49% | 23.63% | 50.73% |
| Retrieval-baseline: classifier head | — | 33.84% | 60.92% | 35.94% | 64.23% | 23.13% | 48.50% |
| Zero-shot-prompting | Llama 3.1 base | 6.97% | 24.41% | 8.04% | 31.86% | 2.77% | 17.30% |
| Zero-shot-prompting | Llama 3.1 PEFT | 26.63% | 58.40% | 33.59% | 65.36% | 21.55% | 56.86% |
| Zero-shot-prompting | Llama 3.3 base | 26.19% | 71.39% | 29.35% | 74.79% | 15.66% | 52.67% |
| Zero-shot-prompting | Llama 3.3 PEFT | 39.92% | 67.98% | 47.85% | 74.25% | 30.34% | 66.15% |
| Three-shot-prompting | Llama 3.1 base | 12.59% | 49.37% | 8.29% | 36.78% | 4.03% | 20.01% |
| Three-shot-prompting | Llama 3.1 PEFT | 24.26% | 52.67% | 31.68% | 59.24% | 19.61% | 47.13% |
| Three-shot-prompting | Llama 3.3 base | 26.19% | 67.79% | 28.82% | 73.52% | 16.68% | 56.98% |
| Three-shot-prompting | Llama 3.3 PEFT | 38.34% | 62.15% | 46.33% | 69.76% | 30.61% | 63.78% |

| | | | | | | | |
|---|---|---|---|---|---|---|---|
| RAG-prompting (similarity search) | Llama 3.1 base | 20.06% | 47.08% | 23.22% | 54.38% | 17.14% | 34.59% |
| RAG-prompting (similarity search) | Llama 3.1 PEFT | 21.39% | 61.51% | 26.10% | 66.57% | 17.78% | 48.55% |
| RAG-prompting (similarity search) | Llama 3.3 base | 39.58% | 74.95% | 44.04% | 78.96% | 27.01% | 63.98% |
| RAG-prompting (similarity search) | Llama 3.3 PEFT | 42.84% | 66.16% | 50.17% | 73.25% | 30.54% | 62.99% |
| RAG-prompting (classifier head) | Llama 3.1 base | 18.92% | 46.84% | 21.23% | 54.39% | 16.25% | 36.53% |
| RAG-prompting (classifier head) | Llama 3.1 PEFT | 20.95% | 59.24% | 25.93% | 64.50% | 16.98% | 49.96% |
| RAG-prompting (classifier head) | Llama 3.3 base | 34.54% | 72.53% | 39.25% | 77.48% | 26.55% | 64.50% |
| RAG-prompting (classifier head) | Llama 3.3 PEFT | 39.48% | 63.88% | 47.31% | 71.37% | 28.95% | 61.46% |

**Table S5** Average total and per-sample inference times of Llama 3.1 8B and Llama 3.3 70B models across ICD-10 and ICD-O code generation tasks

| Model | Quantization | ICD Code | Total Inference time [min] | Inference time per sample [s] |
|---|---|---|---|---|
| Llama 3.1 8B base | No | ICD-10 | 8.76 | 0.23 |
| Llama 3.1 8B PEFT | No | ICD-10 | 8.37 | 0.25 |
| Llama 3.3 70B base | 4-bit | ICD-10 | 35.01 | 1.04 |
| Llama 3.3 70B PEFT | 4-bit | ICD-10 | 28.75 | 0.85 |
| Llama 3.1 8B base | No | ICD-O | 9.39 | 0.28 |
| Llama 3.1 8B PEFT | No | ICD-O | 9.51 | 0.28 |
| Llama 3.3 70B base | 4-bit | ICD-O | 22.70 | 0.67 |
| Llama 3.3 70B PEFT | 4-bit | ICD-O | 20.87 | 0.62 |

**Table S6** Few-shot prompting example illustrating the prompt structure for ICD-10 coding. The texts and codes used there were retrieved from the ICD-10-GM classification.

| German original | English translation |
|---|---|
| **User:** Antworte nur mit dem ICD-10-Code. Was ist der ICD-10-Code für die Tumordiagnose „Bösartiges Melanom des Ohres und des äußeren Gehörganges“? | **User:** Answer only with the ICD-10 code. What is the ICD-10 code for the tumor diagnosis “Malignant melanoma of the ear and external auditory canal”? |
| **Assistant:** C43.2 | **Assistant:** C43.2 |
| **User:** Antworte nur mit dem ICD-10-Code. Was ist der ICD-10-Code für die Tumordiagnose „Carcinoma in situ: Haut des Ohres und des äußeren Gehörganges“? | **User:** Answer only with the ICD-10 code. What is the ICD-10 code for the tumor diagnosis “Carcinoma in situ: skin of the ear and external auditory canal”? |
| **Assistant:** D04.2 | **Assistant:** D04.2 |
| **User:** Antworte nur mit dem ICD-10-Code. Was ist der ICD-10-Code für die Tumordiagnose „Sonstige bösartige Neubildungen: Haut des Ohres und des äußeren Gehörganges“? | **User:** Answer only with the ICD-10 code. What is the ICD-10 code for the tumor diagnosis<br>“Other malignant neoplasms: skin of the ear and external auditory canal”? |
| **Assistant:** C44.2 | **Assistant:** C44.2 |
| **User:** Antworte nur mit dem ICD-10-Code. Was ist der ICD-10-Code für die Tumordiagnose „Spinozelluläres Karzinom, Lokalisation: Ohr Tragus links 3,6mm“? | **User:** Answer only with the ICD-10 code. What is the ICD-10 code for the tumor diagnosis<br>“Squamous cell carcinoma, location: left tragus of the ear, 3.6 mm”? |

**Table S7** Few-shot prompting example illustrating the prompt structure for ICD-O coding. The texts and codes used there were retrieved from the ICD-O-LE dataset and derived from the mapping table of the ICD-10-GM and the ICD-O-3 classification.

| German original | English translation |
|---|---|
| **User:** Antworte nur mit dem ICD-O-Topographie-Code. Wie lautet der ICD-O-Code für die Lokalisation zur Tumordiagnose „Bösartiges Melanom des Ohres und des äußeren Gehörganges (Äußeres Ohr)“? | **User:** Answer only with the ICD-O topography code. What is the ICD-O code for the anatomical site associated with the tumor diagnosis “Malignant melanoma of the ear and external auditory canal (external ear)”? |
| **Assistant:** C44.2 | **Assistant:** C44.2 |
| **User:** Antworte nur mit dem ICD-O-Topographie-Code. Wie lautet der ICD-O-Code für die Lokalisation zur Tumordiagnose „Bösartiges Melanom der Haut, nicht näher bezeichnet (Haut ohne nähere Angabe)“? | **User:** Answer only with the ICD-O topography code. What is the ICD-O code for the anatomical site associated with the tumor diagnosis “Malignant melanoma of the skin, not otherwise specified”?) |
| **Assistant:** C44.9 | **Assistant:** C44.9 |
| **User:** Antworte nur mit dem ICD-O-Topographie-Code. Wie lautet der ICD-O-Code für die Lokalisation zur Tumordiagnose „Bösartiges Melanom der oberen Extremität, einschließlich Schulter (Haut der oberen Extremitäten und der Schulter)“? | **User:** Answer only with the ICD-O topography code. What is the ICD-O code for the anatomical site associated with the tumor diagnosis “Malignant melanoma of the upper limb, including shoulder (skin of upper limb and shoulder)”? |
| **Assistant:** C44.6 | **Assistant:** C44.6 |
| **User:** Antworte nur mit dem ICD-O-Topographie-Code. Wie lautet der ICD-O-Code für die Lokalisation zur Tumordiagnose „Superfiziell spreitendes malignes Melanom linkes Schulterblatt“? | **User:** Answer only with the ICD-O topography code. What is the ICD-O code for the anatomical site associated with the tumor diagnosis “Superficial spreading malignant melanoma of the left shoulder blade”? |
